# Social determinants of COVID-19 mortality at the county level

**DOI:** 10.1101/2020.05.03.20089698

**Authors:** Rebecca K. Fielding-Miller, Maria E. Sundaram, Kimberly Brouwer

## Abstract

The United States is currently the global epicenter of the COVID-19 pandemic. Emerging data suggests that social determinants of health may be key drivers of the epidemic, and that minorities, migrants, and essential workers may bear a disproportionate degree of risk. We used publicly accessible datasets to build a series of spatial autoregressive models assessing county level associations between COVID- 19 mortality and (1) Percentage of Non-English speaking households, (2) percentage of individuals engaged in hired farm work, (3) percentage of uninsured individuals under the age of 65, and (3) percentage of individuals living at or below the poverty line. Across all counties (n=2940), counties with more farmworkers, more residents living in poverty, higher density, and more residents over the age of 65 had significantly higher levels of mortality. In urban counties (n=114), only county density was significantly associated with mortality. In non-urban counties (n=2826), counties with more non-English speaking households and more farm workers had significantly higher levels of mortality, as did counties with higher levels of poverty and more residents over the age of 65. More uninsured residents was significantly associated with decreased reported COVID-19 mortality. Individuals who do not speak English, individuals engaged in farm work, and individuals living in poverty may be at heightened risk for COVID-19 mortality in non-urban counties. Mortality among the uninsured may be being systematically undercounted in county and national level surveillance.

## Introduction

A novel coronavirus responsible for COVID-19 respiratory disease is causing a global pandemic which has already resulted in nearly 5 million cases and over 300,000 deaths since early January(1). The United States currently has more cases than any other nation in the world, with over 2 million cases and 113,000 deaths as of June 11, 2020(1). Preliminary data indicates that existing health inequities in the United States are likely linked to COVID-19 morbidity and mortality(2).

Both infectious and non-communicable disease tends to impact marginalized populations at disproportionate rates. While demographically disaggregated data is not currently available at the national level, data from county and state level entities suggest that COVID-19 may follow similar patterns. In the State of California, Latinos make up approximately 39% of the total population but represent just over 53% of total cases(3). Similarly, in New York City, Black/African American and Hispanic residents have significantly higher rates of COVID-19 illness and mortality than white residents, with a nearly doubled risk of mortality for Black/African American residents compared to white residents(4). While more granular data are not yet available to assess which risk factors may be leading to these disparities in morbidity and mortality, journalistic reporting early analyses suggest that language barriers, poor working conditions among essential workers – who are more likely to be immigrants and/or racial/ethnic minorities(5) - and concerns about immigration status may be creating particular risk among racial and ethnic immigrants across the United States(6, 7).

We sought to assess the associations between COVID-19 mortality and immigrant and farm worker population at the county level. We hypothesized that counties with more immigrants and farm workers would report higher COVID-19 mortality, adjusting for poverty, insurance rates, population age, and density at the county level.

## Methods

We built a series of spatial autoregressive models to assess county-level associations between COVID-19 mortality and: (1) Percentage of Non-English speaking households (defined as households in which no one 14 years or older reports speaking English at least “very well”) and (2) percentage of individuals engaged in hired farm work(8) in the county as of 2018. To account for potential confounders, we adjusted our analyses for the percentage of uninsured individuals under the age of 65, percentage of individuals living at or below the poverty line, percentage of residents age 65 or older, and county density, measured as number of residents per square mile.

COVID-19 mortality data was sourced from county public health agencies, aggregated and made publicly available by the New York Times(9). The proportion of households with limited English speaking ability was drawn from the American Community Survey’s (ACS) 2014 5-year estimate, percentages of individuals living below poverty, and percentage of residents over the age of 65 were from 2017 ACS data. The percentage of farmworkers was taken from the US Bureau of Economic Analysis. Percent uninsured was based on the US Census Small Area Health Insurance Estimates (SAHIE) program’s 2018 estimates. Density was measured as number of individuals per square mile, based on US census data.

In addition to hypothesized predictors and potential confounders, we adjusted our models to account for the stage of the local epidemic by including a variable for the number of “high risk” days a county had experienced. We calculated this number by subtracting the total number of days in which a county had been under a shelter in place order from the total number of days since the first reported case.

Counties with 1000 residents or more per square mile were coded as urban, counties with less than 1000 residents per square mile were coded as urban. While there are many ways to classify counties, we chose to use 1000 people per square mile for two reasons. First, the US census uses this cutpoint to designate census blocks as urban vs. non-urban. Second we felt that doing so allowed us to more clearly delineate major metropolitan areas and their associated resources and public health infrastructures from neighboring suburban or exurban counties.

We first built a series of simple linear regression models to assess the bivariate association between number of deaths within a county and our hypothesized predictors, adjusting for days since 1st case and SIP order. We then constructed a spatial contiguity matrix, and checked the assumption that residuals were distributed spatially using a Moran’s I test.

We next built three separate spatial autoregressive models to assess the association between number of deaths and our hypothesized social determinants, adjusting for potential confounders, and fitted the model with a spatial lag of the dependent variable based on our contiguity matrix. Our first model assessed relationships across all counties. We then stratified our analyses to measure the association between mortality and our hypothesized predictors in urban and non-urban counties.

## Results

This analysis encompassed 2,941 counties across all 50 states. As of June 11, 2020, the number of deaths reported in the NY Times aggregated dataset ranged from 0 to 21,436 per county, with a median of 1 an interquartile range (IQR) of 0-8^1^. We classified 115 counties as urban and 2826 counties as non-urban. Deaths in urban counties ranged from 0 – 21436, with a median of 210 and an IQR of 66-62. Deaths in rural counties ranged from 0-844 with a median of 1 and an IQR of 0-6 (table 1).

**Table 1:** Primary predictor and covariates of interest across all counties and stratified by urban and non-urban.

|  | All counties<br>(n=2941) |  | non-urban counties<br>(n=2826) |  | urban counties<br>(n=115) |  |
| --- | --- | --- | --- | --- | --- | --- |
|  | *median* | *IQR* | *median* | *IQR* | *median* | *IQR* |
| *deaths* | 1 | 0-8 | 1 | 0-6 | 210 | 66.0-662.0 |
| *% Farm workers* | 2.2 | 0.9 – 4.6 | 2.4 | 1.0 – 4.8 | 0.1 | 0.0 – 0.1 |
| *% Non-English speakers* | 5.0 | 2.9 – 10.2 | 4.8 | 2.8 – 9.4 | 19.0 | 11.4 – 29.8 |
| *% Residents uninsured* | 10.5 | 7.4 – 14.5 | 10.6 | 7.4 – 14.6 | 8.3 | 5.9 – 12.6 |
| *% Residents in poverty* | 15.1 | 11.4 – 9.5 | 15.2 | 11.5 – 19.7 | 13.0 | 8.9 – 16.7 |
| *Residents per square mile* | 46.9 | 19.9 – 115.0 | 44.7 | 18.8-101.5 | 1754.9 | 1313.4-2715.3 |
| *% Residents Over 65* | 16.3 | 11.0 – 14.5 | 16.5 | 14.1-19.0 | 12.5 | 11.0-14.5 |

<sup>1</sup> Within this dataset, the 5 boroughs/counties of New York are treated as a single entity. We have done the same in these analyses, assigning all 5 counties the values associated with New York County.

**Table 2:** Full spatial regression models for all counties and stratified by urban/rural.

|  | All counties<br>(n=2940) |  |  |  | non-urban counties<br>(n=2826) |  |  |  | urban counties<br>(n=114) |  |  |  |
| --- | --- | --- | --- | --- | --- | --- | --- | --- | --- | --- | --- | --- |
|  | *b direct* | *b indirect* | *b total* | *p-value* | *b direct* | *b indirect* | *b total* | *p-value* | *b direct* | *b indirect* | *b total* | *p-value* |
| % *Farm workers* | 5.29 | 0.95 | 6.24 | 0.003 | 0.57 | 0.16 | 0.74 | 0.02 | 1956.83 | 130.54 | 2087.37 | 0.20 |
| % *Non-English speakers* | 0.13 | 0.03 | 0.15 | 0.83 | 0.49 | 0.14 | 0.62 | <0.001 | 16.04 | 1.07 | 17.11 | 0.21 |
| % *Residents uninsured* | -1.64 | -0.30 | -1.93 | 0.25 | -0.69 | -0.19 | -0.89 | <0.001 | -50.56 | -3.37 | -53.93 | 0.27 |
| % *Residents in poverty* | 3.50 | 0.63 | 4.12 | 0.001 | 0.32 | 0.09 | 0.40 | 0.01 | 59.86 | 3.99 | 63.85 | 0.10 |
| *Residents per square mile* | 0.33 | 0.06 | 0.39 | <0.001 | 0.17 | 0.05 | 0.21 | <0.001 | 0.35 | 0.02 | 0.38 | <0.001 |
| % *Residents Over 65* | 4.20 | 0.75 | 4.95 | 0.01 | 0.61 | 0.17 | 0.78 | 0.002 | 45.82 | 3.06 | 48.87 | 0.50 |

The Moran’s I test was statistically significant at p <0.01 in each simple (non-spatial) regression, with the exception of associations between density and mortality (Moran’s I p = 0.22) and non-English speakers and mortality (Moran’s I p-value = 0.31) in urban counties, indicating a significant spatial pattern to associations between our hypothesized predictors and mortality.

In our fully adjusted model of all counties, the percentage of farm workers in a county, poverty, population density, and the percentage of residents over the age of 65 were all significantly associated with higher levels of COVID-19 mortality. Each additional percentage of people living in poverty was associated with 3.49 additional deaths within that county (p<0.01), and an additional 0.63 deaths in neighboring counties via a ‘spillover’ effect (p <0.05). Overall, each additional percentage increase of individuals living in poverty within a county was associated with 6.24 additional COVID-19 deaths (p = 0.001). Each additional person per square mile was associated with 0.33 additional deaths within a county and 0.06 indirect deaths, for a total of 0.39 additional deaths (p <0.001). Across all counties, each percentage increase in residents over the age of 65 was associated with 4.20 additional deaths within that county (p = 0.01) and an additional 0.75 deaths in neighboring counties (p = 0.023). Across all counties in the United States, each additional percentage increase in farmworkers was associated with 5.29 additional deaths within a county (p = 0.003) and 0.95 additional deaths in neighboring counties (p = 0.02)

In urban counties (n=114), only population density was significantly associated with higher mortality. In these counties, each additional person per square mile was associated with 0.35 additional deaths within that county (p <0.001).

In non-urban counties, all of our hypothesized social determinants were significantly associated with higher levels of mortality. Each increase in the percentage of farmworkers residing in a county was associated with 0.74 additional deaths (p = 0.02). Each additional percentage of non-English speaking households was associated with 0.62 additional deaths (p <0.001), 0.49 deaths within a given county, and 0.14 attributable to ‘spillover’ effects to neighboring counties (p = 0.002). As in the all county and urban models, density, poverty, and the percentage of residents over the age of 65 were all significantly associated with higher mortality. Contrary to our initial hypotheses, the percentage of uninsured individuals was associated with lower reported COVID19 mortality. In rural areas, each increase in the percentage of uninsured individuals was associated with a direct effect of 0.69 fewer deaths within the county (p <0.001) and 0.19 fewer deaths in neighboring counties (p =0.007).

## Discussion

Although we cannot draw conclusions about individual risk profiles, our findings do suggest that that farm work may create unique risk factors and that farmworkers may require additional protections, such as personal protective equipment and/or targeted outreach. Immigrants provide approximately 75% of all farm labor in the United States(8). Among those engaged in crop work specifically, nearly three quarters are migrants and approximately half are undocumented(8). Undocumented status may impede an individual’s willingness or ability to seek healthcare, or their ability to request additional protections from an employer if they worry doing so could result in their own deportation or that of a family member(10). Farm labor is considered essential work, but there are reports of inadequate personal protective equipment and inadequate social distancing guidelines or enforcement(6). There have also been several high profile outbreaks at meat processing facilities across the country. While the farmworker classification that we used in this analysis does not include individuals engaged in meatpacking, the category does include individuals engaged in livestock care. It is reasonable to assume that some of the risk we identify here may be attributable to the fact that counties with high numbers of farmworkers are also likely to have high numbers of workers in animal processing facilities.

The negative association we found between insured status and mortality is a point of concern. The CDC has noted higher than expected numbers of death across the United States throughout April in recent months, suggesting that COVID-19 mortality is potentially higher than what has thus far been captured by state and county level surveillance(11). It is possible that this association represents a gap in testing and linkage to care among the uninsured, and/or a gap in ascertaining deaths due to COVID-19 among uninsured individuals.

## Data Availability

All data is from publicly available sources and cited as such

## Conclusion

COVID-19 mortality appears to be statistically significantly associated with social determinants of health at the county level, and these relationships may be more pronounced in non-urban counties. Individuals who do not speak English, individuals engaged in farm work, and individuals living in poverty may be at heightened risk for COVID-19 mortality in non-urban counties.

## Acknowledgments

This work was supported by the National Institute of Mental Health, grant K01MH112436 and a National Institute on Minority Health and Health Disparity Loan Repayment Contract, L60-MD011114

## Footnotes

1 Within this dataset, the 5 boroughs/counties of New York are treated as a single entity. We have done the same in these analyses, assigning all 5 counties the values associated with New York County.

## Notes

### Competing Interest Statement

The authors have declared no competing interest.

### Funding Statement

This work was supported by an NIH early career grant award K01MH112436 and a Loan Repayment Contract L60-MD011114

### Author Declarations

UC San Diego IRB

